# Health professionals’ perceptions of human resource management practices and associated workplace factors in public hospitals of Gamo Zone, southern Ethiopia: A cross-sectional study

**DOI:** 10.64898/2026.11.12.26367969

**Authors:** Eden Hailegiorgis, Tezerash Bateno, Tesfahun Mekene, Abayneh Tunje

## Abstract

Health workforce shortages in sub-Saharan Africa are compounded by how existing staff are managed, yet little quantitative evidence describes how hospital health professionals in Ethiopia perceive human resource management (HRM) practices or which workplace conditions accompany those perceptions. Drawing on social exchange theory, we assessed health professionals’ perceptions of HRM practices and their associations with workplace “cost” factors (job demands) and “benefit” factors (job resources) in public hospitals of Gamo Zone, southern Ethiopia. We conducted an institution-based cross-sectional survey from 15 February to 15 March 2025 in three of the zone’s six public hospitals, selected by lottery. Randomly sampled health professionals completed a structured questionnaire on tablets. Perception of HRM practices was measured with 28 five-point items covering seven practice domains and summarized as a mean score from 1 to 5; a mean above 3.0 was classified as a favorable perception. Associations were estimated with multivariable linear regression using heteroscedasticity-robust standard errors, adjusting for age, sex, profession and hospital. After removal of 41 duplicate records and 52 ineligible non-health staff, 343 health professionals were analyzed. The mean HRM perception score was 2.81 (standard deviation 0.58), and 29.0% (99/341; 95% confidence interval [CI] 24.5–34.1%) of participants held a favorable perception. Recruitment and training and development were rated least favorably, and selection most favorably. The proportion with a favorable perception ranged from 1.5% at the general hospital to 55.5% at one primary hospital. In the adjusted model (n = 326; R² = 0.46), greater perceived autonomy (β = 0.20 per scale point; 95% CI 0.06 to 0.35) and recognition (β = 0.12; 95% CI 0.02 to 0.22) were independently associated with more favorable perceptions. Skill ambiguity, work–life role conflict, schedule variability and interpersonal conflict were associated with less favorable perceptions in unadjusted analyses, but no cost factor remained independently associated after adjustment (joint test p = 0.459). Fewer than one in three health professionals viewed HRM practices favorably, with large differences between hospitals. Perceived recognition and autonomy, rather than job demands, were the workplace factors most consistently associated with how staff appraised HRM. These findings point to hospital-level management and non-financial job resources as priorities for health workforce governance research and practice in comparable low-resource settings.

## Introduction

An adequate, motivated and well-managed health workforce is a precondition for universal health coverage. The World Health Organization’s *Global Strategy on Human Resources for Health: Workforce 2030* projected a global shortfall of 18 million health workers [1]; a later reassessment revised this to about 10 million by 2030, with the shortage increasingly concentrated in the WHO African Region [2]. Ethiopia illustrates the challenge: its density of physicians, health officers, nurses and midwives was estimated at 0.96 per 1,000 population, well below the threshold associated with the health-related Sustainable Development Goals [3]. Shortages, however, are only part of the problem. Systematic reviews from low- and middle-income countries consistently identify career development, continuing education, management practices and recognition, alongside pay, as central to health worker motivation and retention [4,5]. Human resource management (HRM)—the planning, recruitment, selection, socialization, development, appraisal and career management of staff—is therefore a core health systems lever rather than an administrative afterthought [6,7].

How HRM affects staff depends not only on formal policies but on how employees experience and interpret them. Research on HR attributions and HRM system strength shows that the same practice can elicit different attitudes depending on whether employees perceive it as fair, consistent and intended to support them [8,9]. In Ethiopian public hospitals, HRM is tightly regulated by government, which constrains hospitals’ latitude to tailor practices, and staff report gaps between intended and implemented practices [10,11]. Ethiopian studies have documented moderate or low job satisfaction and motivation among health professionals [12–15], with non-financial HRM tools among the strongest correlates of motivation [13]. Yet health professionals’ perceptions of HRM practices themselves have rarely been measured. The only Ethiopian study we identified, an unpublished master’s thesis from Metekel Zone (n = 211), reported that 55% of health professionals perceived HRM practices as good and that poor training, workforce planning and performance appraisal were associated with poor perceptions [16]. Hospital-level assessments elsewhere have similarly concentrated on managers’ rather than employees’ views [17].

Social exchange theory offers a useful lens for examining these perceptions. It views employment as a reciprocal exchange in which employees weigh what they invest against what they receive from the organization [18,19]. Organizational support theory, which builds on it, shows that favorable treatment—such as recognition and discretion over one’s work— strengthens employees’ beliefs that the organization values them, whereas unfavorable treatment erodes them [20–22]. A parallel distinction between job demands and job resources underpins the job demands–resources model [23]. Following this logic, we grouped workplace conditions into “cost” factors (workload, skill ambiguity, work–life role conflict, organizational constraints, work schedule variability and interpersonal conflict) and “benefit” factors (social support, recognition and autonomy). Whether demands or resources are more closely tied to how health professionals appraise HRM in low-resource hospitals is not known, and the answer matters for management: resources such as recognition and autonomy may be more amenable to hospital-level action than workload, which is driven largely by staffing.

This study therefore aimed (i) to describe health professionals’ perceptions of HRM practices in public hospitals of Gamo Zone, southern Ethiopia, overall, by practice domain and by hospital, and (ii) to examine the associations of workplace cost and benefit factors with these perceptions, independent of individual characteristics and hospital. Both aims were addressed after an audit of the study dataset that identified and removed duplicate and ineligible records.

## Materials and methods

### Study design and setting

We conducted an institution-based cross-sectional study and report it according to the Strengthening the Reporting of Observational Studies in Epidemiology (STROBE) guideline [24] (S1 Checklist). Gamo Zone lies in South Ethiopia Regional State; its administrative center, Arba Minch, is about 505 km south of Addis Ababa. According to the Gamo Zone Health Department, the zone had six public hospitals—Arba Minch General Hospital and five primary hospitals (Dilfana, Selamber, Chencha, Kamba and Gerese)—employing about 990 health professionals, excluding administrative staff, of whom 412 worked at Arba Minch General Hospital. Data were collected from 15 February to 15 March 2025.

### Participants

The source population was all health professionals employed in public hospitals of Gamo Zone. Health professionals were defined as staff with clinical or allied health professional training (physicians, health officers, nurses, midwives, pharmacy, medical laboratory, health informatics, radiology, anesthesia, environmental and public health professionals, and integrated emergency surgical officers). Health professionals who had worked in a selected hospital for at least six months were eligible. Those holding administrative or leadership positions (for example, hospital directors and unit heads) were excluded because they are responsible for implementing the practices being rated. Non-health support and administrative staff were not part of the study population.

### Sample size and sampling

The sample size was calculated with the single population proportion formula, assuming that 55% of health professionals perceive HRM practices favorably [16], a 95% confidence level and a 5.1% margin of error (n = 366). A finite population correction for an estimated source population of about 994 health professionals gave 268, which was inflated by 10% for non-response and multiplied by a design effect of 1.5 to allow for clustering by hospital, giving 446. Sample sizes calculated for the second objective in Epi Info, using workload and recognition as exposures, were smaller (423 and 378), so 446 was adopted. Three of the six hospitals—Arba Minch General Hospital, Dilfana Primary Hospital and Kamba Primary Hospital—were selected by lottery. The sample was allocated among the selected hospitals, and participants were selected from each hospital’s staff list by simple random sampling.

### Measurements

#### Perception of HRM practices (outcome)

Perception of HRM practices was measured with 28 items covering seven HRM domains described by Dessler [25], each with four items: HR planning, recruitment, selection, onboarding and socialization, training and development, performance appraisal, and promotion, transfer and separation (S1 Text). Items were rated from 1 (strongly disagree) to 5 (strongly agree). The overall perception score was the mean of the 28 items (range 1–5), and domain scores were the means of their four items. The continuous score was the primary outcome. For descriptive purposes, a mean score above 3.0—that is, average responses on the agreement side of “neutral”—was classified as a favorable perception. We chose this scale-anchored threshold in preference to the median split used in the original thesis, because a median split classifies about half of any sample as favorable by construction and discards information [26]; the median-split proportion is reported for comparability.

#### Workplace cost and benefit factors (exposures)

Cost factors were workload (4 items), skill ambiguity (3 items), work–life role conflict (3 items), organizational constraints (5 items), work schedule variability (2 items) and interpersonal conflict (3 items). Benefit factors were social support (4 items), recognition (3 items) and autonomy (6 items). The role conflict items asked about conflict between job demands and responsibilities outside work, so we label this construct work–life role conflict. All items used the same five-point agreement scale and were scored so that higher values indicate more of the named construct (greater demand for cost factors, greater resource for benefit factors); scale scores were item means (range 1–5). The original thesis reverse-coded cost items; this changes the sign but not the magnitude of their coefficients.

#### Covariates

Covariates were age (years), sex, profession and hospital. Profession was grouped as nurse, midwife, health officer, physician (general practitioner or specialist) and other health professional; the last group comprised pharmacy, medical laboratory, health informatics (health information technicians, health statistics and medical records officers), public health, health extension, environmental health, radiology, anesthesia, integrated emergency surgery and infection prevention professionals (S1 Table).

### Data collection and quality control

The structured questionnaire was developed by the investigators from published instruments and Ethiopian studies [10,14,16]. It was prepared in English, translated into Amharic and back-translated to check consistency. It was pretested on about 5% of the sample size at Chencha Primary Hospital, which was not a study hospital, from 10 to 15 February 2025, and wording was revised accordingly. The questionnaire was self-administered on tablets using the KoboCollect application. Trained data collectors were present to explain each section, clarify items on request and check completeness.

### Data audit

Before analysis, we audited the exported data file for duplicate records, out-of-range values, missing data and eligibility. Records identical on all 66 variables were treated as duplicates and only the first occurrence was retained. We reviewed the free-text profession field and classified each respondent as a health professional or as non-health support or administrative staff; the latter did not meet the eligibility criteria and were excluded from the primary analysis but retained in a sensitivity analysis.

### Statistical analysis

Participant characteristics and scale scores were summarized with frequencies, means and standard deviations (SD), and HRM perception scores were compared across groups with t-tests or one-way analysis of variance. Internal consistency was assessed with Cronbach’s alpha and corrected item–total correlations [27]. Dimensionality of the 28-item HRM scale was examined from the eigenvalues of the inter-item correlation matrix, and Harman’s single-factor test was used as a limited check for common-method variance [28]. The proportion with a favorable perception is reported with a Wilson 95% confidence interval (CI).

Associations with the HRM perception score were estimated with linear regression. We first fitted unadjusted models and then a single pre-specified multivariable model that included all nine cost and benefit factors, age, sex, profession and hospital. Variables were selected on theoretical grounds rather than by bivariable p-value screening and stepwise elimination, which inflate type I error and bias coefficients [29]. Hospital was entered as a fixed effect because three clusters are too few for random-effects models or cluster-robust standard errors [30]. Because the Breusch–Pagan test indicated heteroscedasticity, all CIs and p-values were based on HC3 heteroscedasticity-consistent standard errors [31]. Multicollinearity was assessed with variance inflation factors (VIF), residuals were inspected, and influential observations were identified with Cook’s distance. Blocks of cost and benefit factors were tested jointly with Wald tests, and their unique contribution to explained variance was estimated as the change in R². Tests were two-sided with α = 0.05; because the nine factors were specified a priori from theory, no adjustment was made for multiple comparisons. Sensitivity analyses comprised (i) conventional ordinary least squares standard errors; (ii) exclusion of influential observations (Cook’s distance > 4/n); (iii) inclusion of non-health support staff; (iv) analysis of the data file as received, with duplicates retained; (v) replication of the original thesis approach (bivariable screening at p < 0.25 followed by backward elimination at p < 0.05); (vi) logistic regression of favorable perception; and (vii) a recognition-by-hospital interaction. Scale scores were computed when no more than one item was missing for scales of four or more items and when no item was missing for shorter scales; the HRM score required at least 24 of 28 items. Regression analyses used complete cases. Analyses were performed in Python (version 3.13) with the statsmodels package [32]; the analysis code is provided in S1 Code.

### Ethics statement

The study was approved, before data collection began, by the Institutional Research Ethics Review Board of the College of Medicine and Health Sciences, Arba Minch University (reference number IRB/23311/25; protocol number TB23311; approved on 9 January 2025). Letters of permission were submitted to the Gamo Zone Health Department and to each hospital before data collection. Participants were recruited from 15 February 2025 to 15 March 2025. All participants were adult health professionals; each received written information about the study and provided written informed consent before data collection. Names and other direct identifiers were not collected, participation was voluntary, and participants could withdraw at any time. The study was conducted in accordance with the Declaration of Helsinki.

## Results

### Participants

The exported data file contained 436 records. After removal of 41 exact duplicates, 395 unique respondents remained (88.6% of the 446 sampled); 52 of these were non-health support or administrative staff and were excluded, leaving 343 health professionals for analysis (S1 Table). Their mean age was 35.2 years (SD 4.8); 209 (60.9%) were men; nurses (34.7%) and midwives (23.9%) were the largest professional groups; and 39.9% worked at Arba Minch General Hospital (Table 1). Two participants had too many missing HRM items for an outcome score, and 326 (95.0%) had complete data for the multivariable model.

**Table 1.** Characteristics of participating health professionals and HRM perception scores, Gamo Zone public hospitals, 2025 (n = 343).

| **Characteristic** | **Category** | **n (%)** | **HRM perception score, mean (SD)** | **Favorable perception, n/N (%)** | **p-value<sup>c</sup>** |
|---|---|---|---|---|---|
| Sex |  |  |  |  | 0.192 |
|  | Male | 209 (60.9) | 2.84 (0.61) | 69/207 (33.3) |  |
|  | Female | 134 (39.1) | 2.76 (0.54) | 30/134 (22.4) |  |
| Age group, years<sup>b</sup> |  |  |  |  | 0.003 |
|  | <30 | 43 (12.5) | 2.66 (0.45) | 10/42 (23.8) |  |
|  | 30–34 | 99 (28.9) | 2.90 (0.65) | 38/98 (38.8) |  |
|  | 35–39 | 139 (40.5) | 2.88 (0.61) | 42/139 (30.2) |  |
|  | ≥40 | 61 (17.8) | 2.61 (0.44) | 9/61 (14.8) |  |
| Hospital |  |  |  |  | <0.001 |
|  | Arba Minch General Hospital | 137 (39.9) | 2.45 (0.33) | 2/135 (1.5) |  |
|  | Dilfana Primary Hospital | 119 (34.7) | 3.19 (0.70) | 66/119 (55.5) |  |
|  | Kamba Primary Hospital | 87 (25.4) | 2.83 (0.32) | 31/87 (35.6) |  |
| Profession |  |  |  |  | 0.043 |
|  | Nurse | 119 (34.7) | 2.74 (0.51) | 30/119 (25.2) |  |
|  | Midwife | 82 (23.9) | 2.88 (0.69) | 27/82 (32.9) |  |
|  | Health officer | 56 (16.3) | 2.68 (0.53) | 13/54 (24.1) |  |
|  | Physician (general practitioner or specialist) | 26 (7.6) | 2.81 (0.80) | 9/26 (34.6) |  |
|  | Other health professional<sup>a</sup> | 60 (17.5) | 2.97 (0.47) | 20/60 (33.3) |  |
| Total |  | 343 (100) | 2.81 (0.58) | 99/341 (29.0) |  |

HRM, human resource management; SD, standard deviation. HRM perception score is the mean of 28 items scored 1 (strongly disagree) to 5 (strongly agree); favorable perception is a mean score > 3.0; two participants without an outcome score are excluded from the score columns. <sup>a</sup> Pharmacy (n = 16), health informatics (19), medical laboratory (12), public health (4), health extension (3), environmental health (2), and radiology, anesthesia, integrated emergency surgery and infection prevention (1 each). <sup>b</sup> Age missing for one participant. <sup>c</sup> t-test for sex; one-way analysis of variance for other characteristics.

### Measurement properties

All scales showed acceptable to excellent internal consistency (Cronbach’s alpha 0.78–0.91 for the nine workplace factors and 0.79–0.88 for the seven HRM domains; Table 2). The 28-item HRM scale had an alpha of 0.95 and a dominant first factor (first eigenvalue 11.9, explaining 42.5% of variance; second eigenvalue 2.79), supporting the use of an overall score. In Harman’s single-factor test, one factor explained 24.9% of the variance across all 61 items.

**Table 2.** Internal consistency and distribution of workplace factor and HRM perception scales among health professionals (n = 343).

| **Scale** | **Items** | **Cronbach's α** | **Mean (SD)** | **Agree, %** | **Disagree, %** |
|---|---|---|---|---|---|
| **Cost factors (higher = greater demand)** |  |  |  |  |  |
| Workload | 4 | 0.86 | 3.72 (0.60) | 78.7 | 10.8 |
| Skill ambiguity | 3 | 0.89 | 3.53 (0.75) | 68.6 | 18.5 |
| Work–life role conflict | 3 | 0.84 | 3.49 (0.77) | 66.6 | 20.3 |
| Organizational constraints | 5 | 0.86 | 3.61 (0.62) | 71.3 | 13.6 |
| Work schedule variability | 2 | 0.84 | 3.39 (0.80) | 59.9 | 22.8 |
| Interpersonal conflict | 3 | 0.90 | 3.18 (0.90) | 51.5 | 30.2 |
| **Benefit factors (higher = greater resource)** |  |  |  |  |  |
| Social support | 4 | 0.89 | 3.13 (0.81) | 47.7 | 33.7 |
| Recognition | 3 | 0.78 | 2.94 (0.80) | 39.8 | 44.0 |
| Autonomy | 6 | 0.91 | 3.20 (0.77) | 50.8 | 30.2 |
| **Perception of HRM practices** |  |  |  |  |  |
| HR planning | 4 | 0.80 | 2.82 (0.71) | 31.2 | 47.5 |
| Recruitment | 4 | 0.84 | 2.69 (0.70) | 22.5 | 52.0 |
| Selection | 4 | 0.88 | 3.00 (0.78) | 38.8 | 37.9 |
| Onboarding and socialization | 4 | 0.84 | 2.85 (0.76) | 33.4 | 45.7 |
| Training and development | 4 | 0.81 | 2.70 (0.72) | 26.1 | 54.0 |
| Performance appraisal | 4 | 0.79 | 2.78 (0.69) | 29.4 | 48.5 |
| Promotion, transfer and separation | 4 | 0.81 | 2.80 (0.72) | 29.9 | 49.0 |
| Overall HRM perception (28 items) | 28 | 0.95 | 2.81 (0.58) | 30.2 | 47.8 |

HRM, human resource management; SD, standard deviation. Scores are item means (range 1–5). Agree and disagree are the percentages of all item responses that were "agree" or "strongly agree" and "disagree" or "strongly disagree", respectively; the remainder were neutral.

### Perceptions of HRM practices

The mean HRM perception score was 2.81 (SD 0.58; median 2.71, interquartile range 2.43– 3.07). Overall, 99 of 341 health professionals (29.0%; 95% CI 24.5–34.1%) had a favorable perception; using the median split of the original thesis, the figure was 48.7% (95% CI 43.4– 54.0%). Recruitment (mean 2.69) and training and development (2.70) were rated least favorably—more than half of responses in these domains were disagreements—whereas selection was the only domain with a mean at the scale midpoint (3.00) (Fig 1).

**Fig 1.**
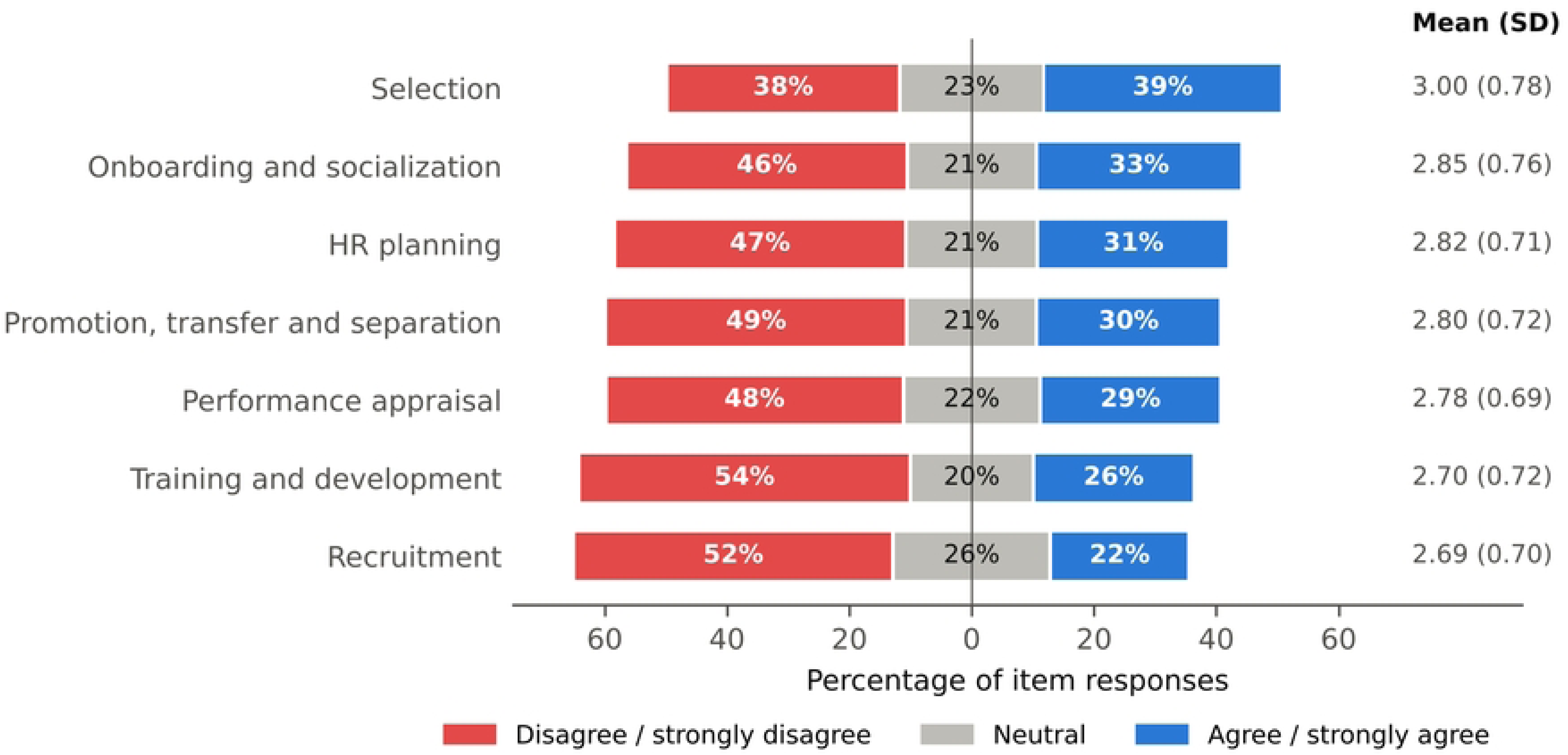
Distribution of responses to HRM perception items by practice domain (n = 343). Bars show the percentage of all item responses in each domain that were disagree or strongly disagree (left), neutral (center) and agree or strongly agree (right); domains are ordered by the proportion agreeing. Values at right are domain mean scores (SD) on a 1–5 scale. HRM, human resource management; SD, standard deviation.

Perceptions varied markedly between hospitals (p < 0.001; Table 1). The mean score was 2.45 at Arba Minch General Hospital, 3.19 at Dilfana Primary Hospital and 2.83 at Kamba Primary Hospital, and the proportion with a favorable perception was 1.5% (2/135), 55.5% (66/119) and 35.6% (31/87), respectively. Hospital alone accounted for 35% of the variance in scores. Scores also differed modestly by profession (p = 0.043) and age group (p = 0.003) but not by sex.

### Workplace cost and benefit factors

Job demands were high. Most item responses indicated agreement with statements describing heavy workload (78.7%), organizational constraints (71.3%), skill ambiguity (68.6%) and work–life role conflict (66.6%) (Table 2). Job resources were rated less favorably: recognition had the lowest mean among the benefit factors (2.94 (0.80)), with more responses disagreeing (44.0%) than agreeing (39.8%) that contributions were recognized.

### Factors associated with perceptions of HRM practices

In unadjusted analyses, all three benefit factors were positively associated with HRM perception scores, and skill ambiguity, work–life role conflict, schedule variability and interpersonal conflict were negatively associated; workload and organizational constraints were not (Table 3; Fig 2). In the multivariable model (n = 326; R² = 0.46; adjusted R² = 0.43), higher autonomy (β = 0.20; 95% CI 0.06 to 0.35; p = 0.006) and recognition (β = 0.12; 95% CI 0.02 to 0.22; p = 0.023) remained independently associated with more favorable perceptions. A one-SD higher autonomy score corresponded to a 0.27-SD higher HRM perception score, and a one-SD higher recognition score to a 0.16-SD higher score. The association with social support attenuated and was no longer statistically significant (β = 0.08; 95% CI −0.05 to 0.20; p = 0.250). None of the cost factors was independently associated with HRM perception (all p ≥ 0.17; joint Wald test p = 0.459), whereas the benefit factors were jointly associated (p < 0.001). The benefit factors uniquely explained 8.0% of the variance in perception scores, compared with 1.8% for the cost factors. After adjustment, scores remained higher at Dilfana than at Arba Minch General Hospital (β = 0.44; 95% CI 0.21 to 0.67; p < 0.001), whereas the unadjusted difference between Kamba and Arba Minch General Hospital disappeared (β = −0.02; 95% CI −0.27 to 0.23; p = 0.865). Age, sex and profession were not independently associated with perceptions.

**Table 3.** Unadjusted and adjusted linear regression of HRM perception score on workplace cost and benefit factors and covariates among health professionals in Gamo Zone public hospitals.

| **Variable** | **Unadjusted β (95% CI)** | **p** | **Adjusted β (95% CI)** | **p** | **Std. β<sup>a</sup>** |
|---|---|---|---|---|---|
| **Cost factors** |  |  |  |  |  |
| Workload | −0.05 (−0.16 to 0.06) | 0.362 | 0.10 (−0.04 to 0.23) | 0.171 | 0.09 |
| Skill ambiguity | −0.17 (−0.26 to −0.07) | <0.001 | −0.01 (−0.13 to 0.11) | 0.887 | −0.01 |
| Work–life role conflict | −0.19 (−0.28 to −0.10) | <0.001 | −0.08 (−0.20 to 0.04) | 0.177 | −0.10 |
| Organizational constraints | −0.08 (−0.19 to 0.03) | 0.163 | 0.01 (−0.10 to 0.13) | 0.820 | 0.01 |
| Work schedule variability | −0.10 (−0.18 to −0.01) | 0.030 | −0.02 (−0.11 to 0.08) | 0.729 | −0.02 |
| Interpersonal conflict | −0.15 (−0.23 to −0.07) | <0.001 | −0.06 (−0.14 to 0.03) | 0.207 | −0.08 |
| **Benefit factors** |  |  |  |  |  |
| Social support | 0.29 (0.21 to 0.36) | <0.001 | 0.08 (−0.05 to 0.20) | 0.250 | 0.10 |
| Recognition | 0.36 (0.28 to 0.43) | <0.001 | 0.12 (0.02 to 0.22) | 0.023 | 0.16 |
| Autonomy | 0.33 (0.25 to 0.41) | <0.001 | 0.20 (0.06 to 0.35) | 0.006 | 0.27 |
| **Covariates** |  |  |  |  |  |
| Age (per year) | −0.008 (−0.018 to 0.002) | 0.100 | −0.003 (−0.013 to 0.007) | 0.539 | −0.03 |
| Female (ref: male) | −0.08 (−0.21 to 0.04) | 0.181 | −0.02 (−0.13 to 0.09) | 0.773 | – |
| **Hospital (ref: Arba Minch General)** |  |  |  |  |  |
| Dilfana Primary Hospital | 0.73 (0.59 to 0.87) | <0.001 | 0.44 (0.21 to 0.67) | <0.001 | – |
| Kamba Primary Hospital | 0.38 (0.29 to 0.47) | <0.001 | −0.02 (−0.27 to 0.23) | 0.865 | – |
| **Profession (ref: nurse)** |  |  |  |  |  |
| Midwife | 0.14 (−0.03 to 0.32) | 0.114 | −0.05 (−0.19 to 0.09) | 0.507 | – |
| Health officer | −0.05 (−0.22 to 0.12) | 0.555 | −0.05 (−0.21 to 0.11) | 0.551 | – |
| Physician | 0.08 (−0.25 to 0.40) | 0.650 | 0.00 (−0.21 to 0.22) | 0.972 | – |
| Other health professional | 0.23 (0.08 to 0.38) | 0.003 | −0.14 (−0.31 to 0.03) | 0.109 | – |

CI, confidence interval; HRM, human resource management. β is the difference in mean HRM perception score (1–5) per one-point higher factor score (1–5), per year of age, or relative to the reference category. All CIs and p-values use HC3 heteroscedasticity-robust standard errors. Unadjusted estimates use all available observations (n = 337–341); the adjusted model (n = 326) includes all variables shown; R² = 0.46, adjusted R² = 0.43, F(17, 308) = 15.7. <sup>a</sup> Standardized β, shown for continuous variables only. Full model output is provided in S3 Table.

**Fig 2.**
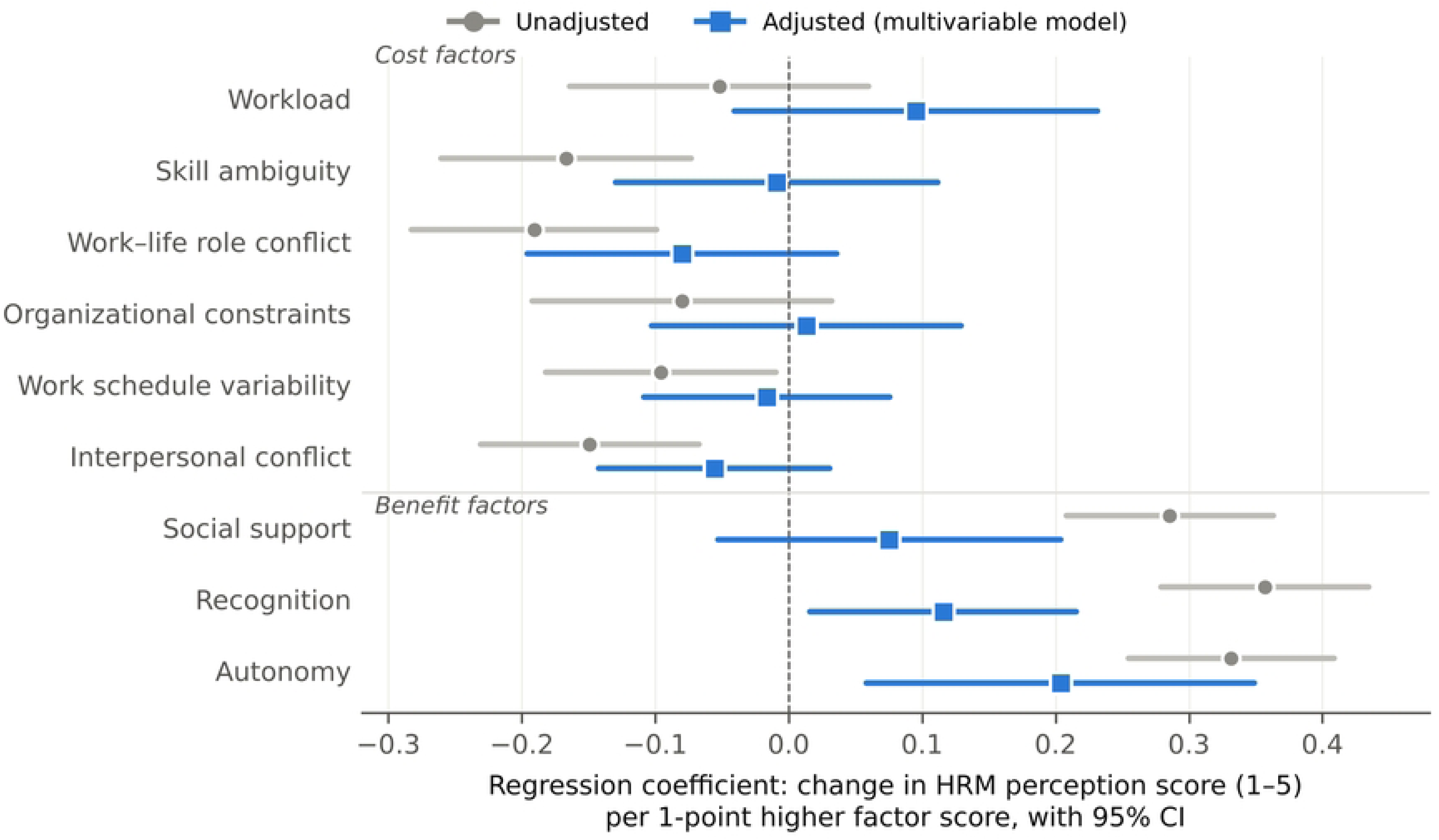
Unadjusted and adjusted associations of workplace cost and benefit factors with HRM perception scores. Points show regression coefficients (change in mean HRM perception score on a 1–5 scale per one-point higher factor score) and lines their 95% confidence intervals based on HC3 heteroscedasticity-robust standard errors. Adjusted estimates are from a single linear regression model including all nine factors, age, sex, profession and hospital (n = 326). HRM, human resource management.

Model diagnostics did not indicate problematic multicollinearity (maximum VIF 4.39, for hospital; all workplace factors < 2.8). Residuals were heteroscedastic (Breusch–Pagan p < 0.001), which the robust standard errors address; 34 observations had a Cook’s distance above 4/n, but none exceeded 0.054.

### Sensitivity analyses

The associations of autonomy and recognition were consistent across all model specifications, including analyses that added non-health support staff or retained duplicate records (β 0.20–0.23 for autonomy and 0.10–0.16 for recognition; S3 Table). Excluding influential observations strengthened the associations of social support, skill ambiguity and interpersonal conflict, so the null findings for individual cost factors should be regarded as imprecise rather than as evidence of no association. Replicating the original thesis approach of bivariable screening and backward elimination retained hospital, interpersonal conflict, recognition and autonomy. In logistic regression of favorable perception, the odds ratios for autonomy (1.84; 95% CI 0.95–3.54) and recognition (1.56; 95% CI 0.98–2.49) were in the same direction but imprecise, as expected after dichotomizing the outcome. There was weak evidence that the association with recognition differed by hospital (interaction p = 0.073).

## Discussion

In this survey of health professionals in three public hospitals in southern Ethiopia, fewer than one in three held a favorable view of HRM practices, and perceptions of recruitment and of training and development were particularly negative. Perceptions varied more between hospitals than across any individual characteristic. Among workplace factors, recognition and autonomy—two job resources—were independently associated with more favorable perceptions, whereas job demands such as workload, skill ambiguity and work–life role conflict, although high, were not independently associated once resources and hospital were taken into account.

The proportion with a favorable perception (29.0%) was lower than the 55% reported in Metekel Zone [16]. The domain pattern is more informative and is consistent with prior Ethiopian evidence. In Metekel, poor training and development, workforce planning and performance appraisal were associated with poor HRM perceptions [16], and a meta-analysis of Ethiopian studies identified opportunities for professional growth as a strong correlate of job satisfaction [12]. Qualitative work in Ethiopian public hospitals describes centrally regulated staffing, salaries and employment arrangements that leave hospitals little room to tailor recruitment and development and that leave staff doubting management’s ability to respond to workforce needs [10,11]. Our finding that recruitment was among the least favorably rated domains fits this description.

The large between-hospital differences—from 1.5% favorable at the general hospital to 55.5% at Dilfana Primary Hospital—suggest that perceptions of HRM are partly a property of the organization rather than of the individual. This is consistent with the notion of HRM system strength, in which the consistency and visibility of management practices shape shared employee perceptions [9], and with Ethiopian evidence that health worker motivation varies between hospitals [14]. With only three hospitals, however, we cannot identify which hospital characteristics—size and patient load, leadership, staff composition or local implementation of national policies—explain these differences. The persistence of the difference between Dilfana and Arba Minch General Hospital after adjustment indicates that it was not explained by the individual-level factors we measured.

The independent associations of recognition and autonomy with HRM perceptions are consistent with social exchange and organizational support theories, which predict that discretionary favorable treatment signals that the organization values its employees [19–21], and with job design research identifying autonomy as a core motivating job characteristic [33]. They also echo reviews and Ethiopian studies in which recognition and other non-financial HRM tools were among the most influential determinants of health worker motivation [4,5,13]. Alternative explanations must be considered. Because all measures were self-reported at one time point, the associations may reflect reverse causation—staff who view HRM favorably may also report more recognition—or a general evaluative disposition that colors responses to all items.

### Strengths and limitations

Strengths of this study include random selection of hospitals and participants, a high response proportion, a theory-guided and pre-specified analysis, scales with good internal consistency, robust standard errors and extensive sensitivity analyses. We also audited the data before analysis and report how duplicate and ineligible records were handled.

Limitations should also be considered. First, the cross-sectional design precludes causal inference, and the direction of associations cannot be established. Second, all variables were self-reported, so shared method variance and social desirability may have inflated associations. Third, important covariates such as years of service, education, salary, employment status and department were not collected, so residual confounding is likely.

Finally, the study included three hospitals in one zone; hospital effects are confounded with unmeasured hospital characteristics, and the findings may not generalize to other regions, to referral hospitals or to primary health care facilities.

### Implications

For hospital managers and zonal health departments, the findings suggest two practical priorities. First, because perceptions differed so much between hospitals, hospital-level diagnosis of HRM practices—particularly recruitment, training and development, and performance appraisal—may be more useful than uniform zonal responses. Second, recognition and decision latitude are non-financial resources that are at least partly within managers’ control even under centrally regulated pay and staffing. Whether strengthening them improves perceptions of HRM, and ultimately motivation, retention or quality of care, needs to be tested in longitudinal or intervention studies that use validated instruments, include more hospitals so that organizational factors can be modeled, and measure outcomes that matter for patients.

## Conclusions

Fewer than one in three health professionals in public hospitals of Gamo Zone perceived HRM practices favorably, with recruitment and training and development rated least favorably and large differences between hospitals. Perceived recognition and autonomy were independently associated with more favorable perceptions, whereas job demands were not after adjustment. These cross-sectional associations identify hospital-level management and non-financial job resources as promising targets for further research and for efforts to strengthen health workforce governance in low-resource settings.

## Data Availability

There is data and will be available on request.

## Acknowledgments

We thank the Gamo Zone Health Department and Plan Department; the management and staff of Arba Minch General Hospital, Dilfana Primary Hospital and Kamba Primary Hospital; the data collectors; and the health professionals who participated in the study.

## Supporting information

**S1 Checklist. STROBE checklist for cross-sectional studies.**

**S1 Table. Participant flow, data audit and classification of professions.** Duplicate records removed, non-health support staff excluded and the professions grouped as “other health professional”.

**S2 Table. Item-level responses to all questionnaire items (n = 343).** Frequencies of each response option, missing responses and item means.

**S3 Table. Full multivariable regression output and sensitivity analyses.** Coefficients, standard errors, t-statistics, p-values and confidence intervals (conventional and HC3), model fit, and estimates from alternative model specifications.

**S1 Text. Study questionnaire (English version).**

**S1 Code. Python code used for the data audit and analyses.**

**S1 Dataset. De-identified participant-level dataset and data dictionary.**

## Notes

### Competing Interest Statement

The authors have declared no competing interest.

### Author Declarations

The study was approved, before data collection began, by the Institutional Research Ethics Review Board of the College of Medicine and Health Sciences, Arba Minch University (reference number IRB/23311/25 protocol number TB23311 approved on 9 January 2025). Letters of permission were submitted to the Gamo Zone Health Department and to each hospital before data collection. Participants were recruited from 15 February 2025 to 15 March 2025. All participants were adult health professionals each received written information about the study and provided written informed consent before data collection. Names and other direct identifiers were not collected, participation was voluntary, and participants could withdraw at any time. The study was conducted in accordance with the Declaration of Helsinki.

